# Age-varying genetic associations and implications for bias in Mendelian randomization

**DOI:** 10.1101/2021.04.28.21256235

**Authors:** Jeremy A Labrecque, Sonja A Swanson

## Abstract

Estimates from conventional Mendelian randomization (MR) analyses can be biased when the genetic variants proposed as instruments vary over age in their relationship with the exposure. For four exposures commonly studied using MR, we assessed the degree to which their relationship with genetic variants commonly used as instruments varies by age using flexible, spline-based models in UK Biobank data. Using these models, we then estimated how biased MR estimates would be due to age-varying relationships using plasmode simulations. We found that most genetic variants had age-varying relationships with the exposure for which they are a proposed instrument. Body mass index and LDL cholesterol had the most variation while alcohol consumption had very little. This variation over age led to small potential biases in some cases (*e*.*g*. alcohol consumption and C-reactive protein) and large potential biases for many proposed instruments for BMI and LDL.

## Introduction

Conventional Mendelian randomization (MR) is an application of instrumental variable analysis with genetic variants proposed as instruments. To be an instrument, a genetic variant must be associated with the exposure, must only affect the outcome through its effect on the exposure and must not be confounded with the outcome.^1^ Many, if not most, non-MR applications of instrumental variable analysis propose instruments that operate on a short time scale, affecting the exposure at a single time point or for a window of time on the order of perhaps days, weeks, or months. In MR, there is often a decades-long period of time between when the proposed instrument is set (at conception) and when the exposure and outcome are measured.^2^ This allows MR estimates to be interpreted as “lifetime effects” because the proposed genetic instrument affects or is correlated with the exposure’s trajectory across the lifetime.^3,4^ We have previously shown that such an interpretation requires more than the core MR assumptions^3^: to use conventional MR methods requires another assumption such as that the genetic variant’s relationship with the exposure must also stay constant over time.

Genetic epidemiologists have identified genetic variants whose relationship with a given phenotype varies with age^5–7^, including for genetic variants associated with body mass index (BMI) and low density lipoprotein cholesterol (LDL). Nearly all exposures used in MR are age-varying, i.e. the exposure can vary with age. For most combinations of genetic variants and exposures used in MR, however, it is not known the degree to which the associations between these genetic variants and exposure vary with age. Thus, the degree to which variations in these genetic associations would cause bias is also not known. If age-varying relationships between proposed genetic instruments and exposures is common and causes important bias in conventional MR approaches, it would be invaluable information for investigators considering conducting MR studies. Such information may help them exclude variants that would clearly lead to biases, better understand the magnitude or direction of bias in conventional MR methods, or seek out alternative MR methods when repeated exposure measures are available.

Our first objective is to examine the age-varying association between genetic variants often used in MR studies and the exposures for which they are proposed instruments in the UK Biobank. Specifically, we estimate the association between genetic variants and four exposures: body mass index (BMI), alcohol consumption, C-reactive protein (CRP) and low-density lipoprotein cholesterol (LDL). After estimating age-varying relationships between genetic variants and their associated exposures, our second objective is to estimate the possible bias in conventional MR estimates due to these age-varying associations seen in our first objective. To do this, we will use the observed relationships from the first objective while simulating an outcome with a known relationship to the exposure. By estimating the effect of the exposure using MR, we can describe how much bias can result from naturally occurring age-varying genetic association.

## Methods

### Data source

We used data from the UK Biobank, which recruited 503,325 participants from 22 assessment centers in the United Kingdom between 2006 and 2010.^8^ To minimize population stratification, we restricted the sample to unrelated, white participants of British ancestry. Questionnaire data, physical measures and biological samples were taken at recruitment. The UK Biobank is often used for MR studies and has a large number of participants from a wide range of ages, 40 to 70 years at baseline, over which to assess age-varying genetic associations. We excluded the few participants below age 40. The UK Biobank has ethics approval from the UK National Health Service’s National Research Ethics Service (ref 11/NW/0382).

### Exposures and genetic variants

We selected four exposures that are commonly used in MR studies in order to explore to what extent genetic variants proposed as instrumental variables for these exposures vary by age. We did not select the exposures with the goal of generalizing to other exposures because age-varying genetic relationships are likely to differ depending on the exposure. Instead, we selected two genetically distal (BMI, alcohol consumption) exposures and two genetically proximal (CRP, LDL) to examine a variety of exposure types.

BMI was calculated from the height measured using a Seca 202 device and weight measured with light clothing and without shoes using a Tanita BC418MA body composition analyzer. Alcohol servings per week was measured by combining the self-reported weekly intake of white wine, red wine, beer/cider, spirits and fortified wine.^9^ CRP was measured from a blood sample using a immunoturbidimetric method (high sensitivity analysis on a Beckman Coulter AU5800). LDL was also measured from a blood sample using and enzymatic protective selection analysis on a Beckman Coulter AU5800. The age of participants used in the analysis was their age on the day they attended the assessment center for their baseline measurements.

For each exposure, we chose a set of genetic variants that were previously proposed as instruments in prior MR studies. For alcohol consumption and C-reactive protein, only a small set of genetic variants are commonly used as instruments. We selected genetic variants that appeared in at least five MR studies over the past 10 years. Though rs671 is commonly used for MR studies of the effect of alcohol, it was nearly monogenic in our sample and we therefore excluded it. For BMI, and LDL cholesterol, a larger number of genetic variants are typically used in MR studies. We chose the ten genetic variants with the lowest p-values from the most recent genome-wide association study of each.^10,11^ We also generated a polygenic risk score for BMI and LDL cholesterol using the coefficients reported in the genome-wide association studies, as this is a common practice for MR studies using many variants. All genetic variants were coded so an increase in the number of copies of an allele or score corresponded with an increase in the exposure.

### Estimating age-varying genetic associations

We analyzed each genetic variant-exposure pair (or polygenic risk score-exposure pair) with separate linear regression models. Age was modeled with cubic B-splines with 3 internal knots using the mgcv package^12^ in R.^13^ An interaction between age and number of copies of allele was included to allow the relationship between age and the exposure to be differ in participants with 0, 1 and 2 copies of each genetic variant on the additive scale. The association between a one-unit increase in the polygenic risk scores and the exposure was modeled linearly but this association was also allowed to vary across ages with an interaction between the score and age. All models were also adjusted for the first 10 genetic principal components. We estimated the additive per allele effect as a function of age and its standard error using 1000 samples of the posterior distribution from the linear regression model.^12^

### Plasmode simulations

Previous work has shown that when the relationship between the genetic variant and the exposure varies with age, the conventional MR estimate can be a biased estimate of the lifetime average effect.^3^ To obtain an unbiased lifetime effect with conventional MR methods, further assumptions are needed about either the variant-exposure association or exposure-outcome relationships. For example, if the variant-exposure association at the time the exposure is measured is equal to the variant-exposure association during the exposure window (i.e., the time period during which the exposure has a direct causal effect on the outcome), then along with the usual assumptions for IV estimation we can identify the lifetime average causal effect. If the association varies with age, it is likely that these two values will differ, which would likely bias the MR estimate.

Therefore, we used plasmode simulation to estimate how age-varying effects could potentially bias MR estimates. A plasmode simulation uses an observed data set but simulates one or more variables with the goal of controlling or setting important parameters.^14^ This can be useful to test a method under relatively realistic conditions. Here, we used UK Biobank data for the genetic variant, exposure and principal components but a simulated outcome for which the true lifetime average causal effect was known.

For each genetic variant or score-exposure pair we used the model from the previous section to simulate the exposure trajectory for every participant from age 40 to 65 given their observed genetic variants. Of note, this means we are using cross-sectional data to infer individual-level trajectories over age; we return to this point in the discussion. We then simulate a continuous outcome at age 65 as a function of the exposure trajectory and a hypothetical exposure window. The exposure window defines the magnitude of the effect of the exposure on the outcome at different ages. Exposure windows of different lengths (5, 10, 25 years) and shapes (linear and Gaussian) were used (Figure 1) to examine how the bias changed with the type of exposure window. The outcome was simulated on the additive scale by multiplying the exposure value by the value of the exposure window at every age and summing the result. In other words, we are using a discrete approximation of taking the area under the curve that results from multiplying the density of the exposure window by the value of the exposure. For example, the outcome using a 5 year linear window would be calculated by multiplying the exposure window by the predicted level of the exposure from age 61 to 65 (5 years) and summed. In this way, for this exposure window, the exposure closer to age 65 has a larger effect on the outcome than the exposure earlier in the exposure window. The exposure windows were scaled to make the causal effect of increasing the exposure trajectory by one unit equal to 1. Thus, the simulated outcome was generated so as not to violate the instrumental variable assumptions, *i*.*e*. the outcome shared no common cause with the genetic variants, was only caused by a genetic variant through its effect on the exposure and was homogeneous across all participants.This mimics a data set where the outcome was measured in all participants at age 65.

**Figure 1:**
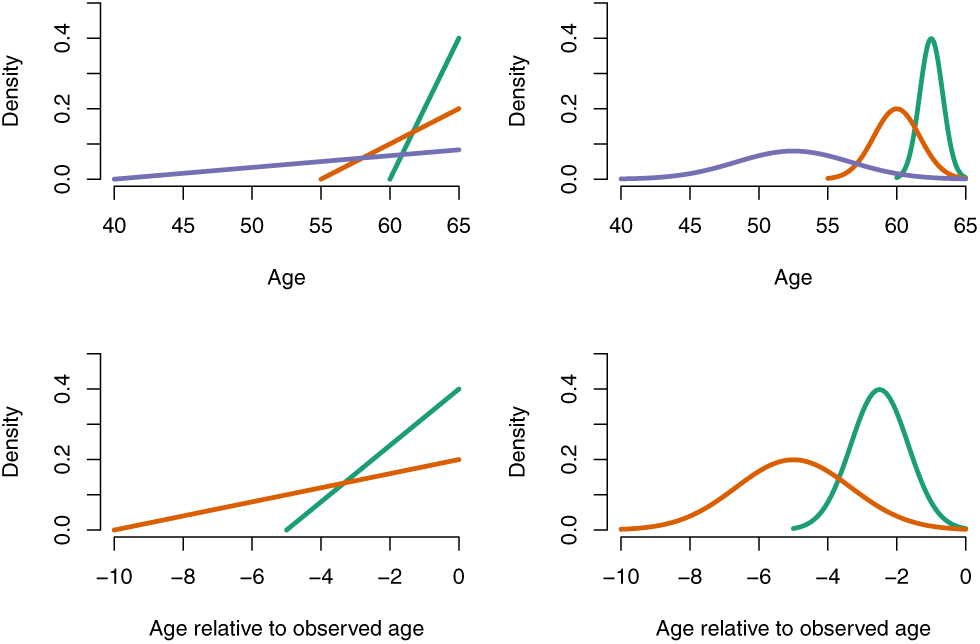
Top left: The three linear exposure windows considered for 5 years (green), 10 years (orange) and 25 years (purple). Top right: The gaussian exposure windows, 5-year window centred at 62.5 years (green), 10-year window centered at 60 years (orange) and 25-year window centered at 52.5 years (purple). Bottom left: The two linear exposure windows relative to observed age for 5 years (green) and 10 years (orange). Bottom right: The two gaussian exposure windows relative to observed age centred at observed age-2.5 (green) and observed age-5 (orange).

Separately, a second outcome was simulated at the age at which each participant was observed at baseline in the UK Biobank. This mimics a data set where all outcomes were measured at baseline. Exposure windows were not defined by age as in the previous paragraph but by time since exposure, thus representing a different biological process. For example, a five-year exposure window for a participant who was 45 years old at baseline was applied from age 41 to 45. Similarly, the same exposure window for a participant who was 58 years old at baseline would be applied from age 54 to 58. The 25 year exposure window was not used for this outcome because it could only include participants at age 65 and would have been equivalent to the 25 year exposure window from the previous paragraph.Using this data set (composed of both observed and simulated variables), we estimated the causal effect separately using each proposed instrument, for each exposure window and for each outcome. The ratio estimator was used assuming the exposure was measured at the same time as outcome, to simulate a cross-sectional study. All analyses were carried out in R.^13^ Functions to carry out the analyses or to apply the analyses to new data sets can be found at https://github.com/jalabrecque/MRcheck.

## Results

After exclusions, the study population consisted of 355,655 participants. The proportion of missing values was 0.3% for BMI, 29.5% for alcohol consumption, 4.9% for C-reactive protein and 4.8% for LDL. The sample size for each exposure can be found in Table A1.

The overall level of each exposure differed by age (Figure 2 and Supplementary Material). BMI increased to about age 60 stabilizing afterward. Alcohol consumption appeared to increase from around age 40 to 55 and decrease afterward but with wide confidence intervals. CRP increased across the entire age range. LDL increased to around age 55 and decreased afterward.

**Figure 2:**
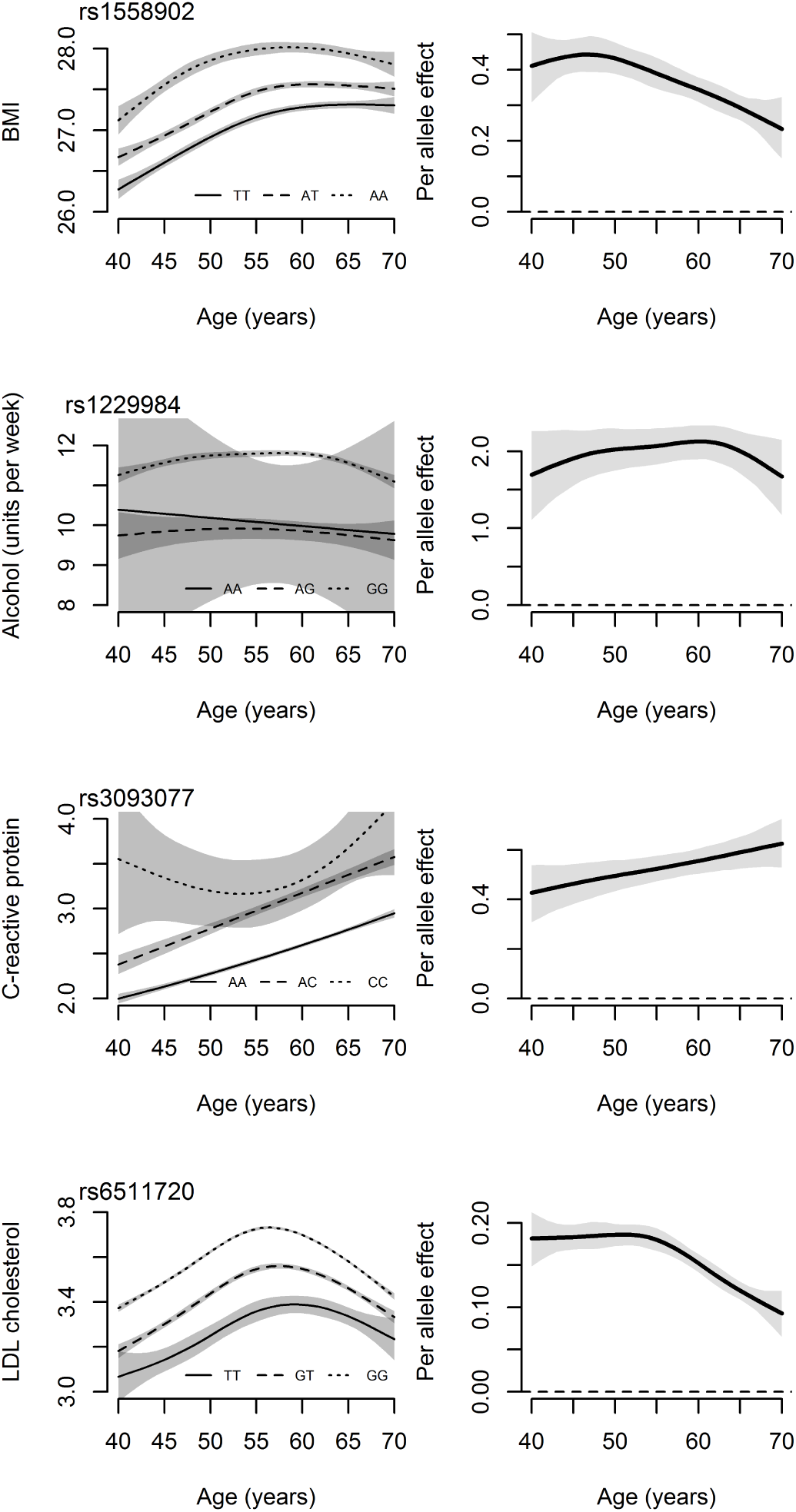
An example of one genetic variant-exposure relationship for each exposure. The left column plots the age-exposure relationship for each genotype of the genetic variant. The right column plots the per allele affect by age.

For most genetic variants, there was evidence of a change in genetic association by age. Figure 2 shows the relationship between age and exposure for different alleles for the genetic variant with the strongest association with an exposure. For each exposure, the trajectory followed by each allele is roughly similar but the per-allele effect changes with age. For BMI and LDL cholesterol, the per allele effect shows an important decrease from age 40 to 70. The per-allele effect for alcohol consumption is relatively stable but with wide confidence intervals. Lastly, for CRP, the per-allele effect increased with age. Graphs for all other genetic variants can be found in the Supplementary Material.

Biases were found in both directions and across a range of magnitudes (Table 1). Large biases were found for both BMI and LDL cholesterol nearing 100% bias for the 25-year Gaussian exposure window. The smallest biases were found for alcohol consumption though the confidence intervals are consistent with important negative and positive biases particularly for rs698 indicating that uncertainty makes it difficult to draw strong conclusions. The biases for genetic variants used as instruments for CRP were all negative and small in magnitude.

**Table 1:** Bias (in percent) for each genetic-variant exposure combination under all exposure window lengtt, exposure window shape and age when the outcome was measured scenario.

|  | At age 65 |  |  |  |  |  | At observed age |  |  |  |
| --- | --- | --- | --- | --- | --- | --- | --- | --- | --- | --- |
|  | Linear exposure window |  |  | Gaussian exposure window |  |  | Linear exposure window |  | Gaussian exposure window |  |
|  | 5 year | 10 year | 25 year | 5 year | 10 year | 25 year | 5 year | 10 year | 5 year | 10 year |
| **BMI** |  |  |  |  |  |  |  |  |  |  |
| rs1558902 | 5 (0,9) | 10 (3,19) | 25 (12,39) | 9 (1,17) | 18 (5,31) | 37 (20,57) | 2 (1,4) | 7 (4,10) | 4 (2,6) | 10 (6,15) |
| rs6567160 | 6 (-1,14) | 10 (-1,24) | 19 (1,43) | 11 (-1,24) | 16 (-2,38) | 26 (1,61) | 2 (0,4) | 4 (-1,9) | 3 (-1,7) | 6 (-2,15) |
| rs13021737 | 15 (6,28) | 33 (15,59) | 65 (31,116) | 29 (12,52) | 55 (25,99) | 89 (41,163) | 4 (2,6) | 10 (4,15) | 7 (3,11) | 15 (6,24) |
| rs10938397 | 11 (1,25) | 15 (-3,38) | 23 (-6,64) | 18 (0,43) | 18 (-10,55) | 32 (-6,87) | 2 (0,5) | 6 (-1,13) | 4 (-1,9) | 9 (-2,22) |
| rs543874 | 6 (-3,18) | 19 (2,42) | 51 (19,101) | 13 (-3,35) | 36 (9,76) | 78 (32,147) | 5 (3,8) | 12 (6,18) | 10 (5,14) | 19 (10,29) |
| rs2207139 | 10 (-1,20) | 13 (-3,32) | 29 (3,66) | 16 (-2,35) | 16 (-8,50) | 45 (7,100) | 2 (-1,4) | 8 (2,16) | 3 (-2,8) | 13 (3,25) |
| rs11030104 | 0 (-9,9) | 2 (-12,22) | 8 (-13,40) | 0 (-15,18) | 6 (-15,38) | 14 (-16,56) | 2 (0,5) | 3 (-4,10) | 4 (-1,9) | 5 (-6,17) |
| rs3101336 | 18 (-2,52) | 39 (1,107) | 79 (13,216) | 34 (-1,95) | 66 (5,179) | 109 (17,301) | 4 (1,8) | 11 (1,22) | 7 (1,15) | 16 (1,35) |
| rs7138803 | 2 (-9,13) | 6 (-11,29) | 19 (-11,65) | 4 (-15,25) | 12 (-16,50) | 30 (-13,96) | 2 (-1,6) | 6 (-2,15) | 4 (-2,10) | 9 (-4,25) |
| rs10182181 | 12 (4,24) | 26 (10,48) | 41 (14,80) | 23 (8,45) | 42 (16,78) | 51 (13,108) | 3 (1,5) | 5 (-1,11) | 5 (1,9) | 7 (-2,17) |
| Score | 4 (3,5) | 9 (7,11) | 23 (18,29) | 7 (6,9) | 15 (11,19) | 36 (28,46) | 3 (2,4) | 7 (6,9) | 5 (4,7) | 12 (9,14) |
| **Alcohol** |  |  |  |  |  |  |  |  |  |  |
| rs1229984 | 3 (-1,6) | 4 (-2,11) | 3 (-8,18) | 5 (-1,11) | 5 (-5,17) | 2 (-13,23) | 0 (-2,1) | -1 (-4,3) | -1 (-4,2) | -2 (-7,5) |
| rs698 | 13 (-6,41) | 28 (-8,97) | 44 (-12,164) | 25 (-10,79) | 45 (-12,160) | 52 (-20,201) | 0 (-4,5) | 3 (-8,14) | 0 (-8,9) | 2 (-14,20) |
| **C-reactive protein** |  |  |  |  |  |  |  |  |  |  |
| rs1130864 | -3 (-6,0) | -4 (-10,2) | -8 (-16,3) | -5 (-11,1) | -5 (-15,4) | -10 (-23,4) | -1 (-2,1) | -2 (-6,1) | -1 (-4,2) | -4 (-9,2) |
| rs1205 | -2 (-4,0) | -2 (-6,2) | -7 (-13,0) | -3 (-6,1) | -3 (-9,3) | -12 (-21,-2) | -1 (-2,0) | -3 (-6,-1) | -2 (-4,0) | -5 (-9,-2) |
| rs3093077 | -1 (-3,0) | -3 (-6,1) | -8 (-16,0) | -3 (-6,1) | -5 (-11,1) | -13 (-24,1) | -2 (-3,0) | -3 (-7,0) | -3 (-6,0) | -5 (-11,0) |
| **LDL cholesterol** |  |  |  |  |  |  |  |  |  |  |
| rs6511720 | 7 (4,10) | 16 (10,22) | 34 (25,45) | 13 (7,19) | 28 (19,38) | 47 (35,63) | 3 (2,4) | 7 (5,9) | 5 (4,6) | 10 (7,14) |
| rs629301 | 3 (0,7) | 11 (5,17) | 30 (19,43) | 7 (1,13) | 22 (12,33) | 46 (30,62) | 3 (3,4) | 8 (5,10) | 6 (5,8) | 12 (9,16) |
| rs1367117 | 5 (1,9) | 11 (4,19) | 27 (13,41) | 9 (3,16) | 20 (7,33) | 39 (21,60) | 3 (1,4) | 7 (4,10) | 5 (2,7) | 11 (6,16) |
| rs4420638 | 4 (1,6) | 10 (5,14) | 23 (15,32) | 8 (3,12) | 18 (10,26) | 34 (23,45) | 2 (1,3) | 5 (3,7) | 4 (2,5) | 8 (5,11) |
| rs4299376 | 7 (0,13) | 13 (3,27) | 23 (4,45) | 13 (0,24) | 20 (4,42) | 32 (7,63) | 3 (1,5) | 5 (1,10) | 6 (2,10) | 9 (1,17) |
| rs2479409 | 7 (-6,26) | 28 (3,70) | 62 (16,140) | 17 (-6,51) | 56 (10,129) | 85 (23,198) | 6 (3,9) | 10 (3,18) | 10 (5,17) | 16 (4,30) |
| rs1564348 | 12 (-6,34) | 36 (5,86) | 68 (14,170) | 25 (-5,68) | 66 (13,155) | 90 (15,228) | 4 (0,8) | 7 (-2,15) | 7 (0,15) | 10 (-4,25) |
| rs11220462 | 7 (-5,20) | 13 (-7,40) | 18 (-10,63) | 12 (-8,37) | 20 (-11,65) | 25 (-14,84) | 3 (0,7) | 3 (-5,11) | 6 (0,13) | 5 (-8,18) |
| rs6029526 | 3 (-9,16) | 7 (-11,32) | 17 (-12,61) | 6 (-15,29) | 13 (-17,57) | 24 (-16,81) | 1 (-2,5) | 4 (-4,13) | 2 (-4,10) | 6 (-7,21) |
| rs635634 | 10 (1,20) | 24 (7,44) | 54 (25,96) | 20 (3,38) | 41 (13,77) | 78 (36,138) | 4 (2,6) | 11 (5,16) | 7 (3,10) | 16 (8,26) |
| Score | 3 (2,4) | 6 (4,8) | 16 (10,22) | 5 (3,7) | 10 (7,14) | 25 (16,35) | 2 (2,3) | 5 (4,7) | 4 (3,5) | 9 (6,11) |

The bias due to age-varying genetic associations were more pronounced when the outcome was simulated at age 65 than when simulated at the observed age. The largest biases for the 10 year window when the outcome was simulated at age 65 were in the range of 50-60% whereas the largest biases for the outcome simulated at the observed age for the same exposure window were between 15-20%. Generally, the bias also increased with the length of the exposure window. This is expected because the per-allele effect changes less in a five year window than it does in a longer 25 year window. Similarly, the bias was larger for the Gaussian exposure window when compared to the linear exposure window. This is also expected because the peak of the linear exposure window occurs simultaneously with the measurement of the exposure whereas the peak for the Gaussian window occurs earlier. The biases estimated using the polygenic risk scores were in the middle range of the biases estimated with the genetic variants individually.

## Discussion

We demonstrated that, for four exposures commonly used in MR studies, the association between genetic variants proposed as instruments and the exposure they proxy varies with age. The patterns of how the association changes with age on the additive scale was different between the four exposures we examined suggesting that patterns in age-varying genetic relationships may be exposure specific.

For many genetic variants seen in this paper, age-varying genetic associations can very well be the cause of important biases in MR estimates even when all the core instrumental variable assumptions were satisfied, as demonstrated in our simulations. Each exposure examined suffered from this bias to a differing degree. The bias for alcohol consumption and CRP were mostly small (except rs698 which had wide confidence intervals) and the biases for BMI and LDL cholesterol were large with magnitudes over 80% in some cases. Within these exposures with high bias, there was a lot of variability between genetic variants though given the wide standard errors of the bias estimates themselves, the differences may be due to random error.

Bias tended to increase with the length of the exposure window. This is to be expected as shorter exposure windows give the relationship between genetic variant and the exposure less time to vary. Therefore, the measured relationship between the genetic variant and the exposure cannot be very different from the average over the short exposure window. This suggests that when studying exposures that only have short term effects on the outcome, the bias due to age-varying genetic associations is likely to be small. However, one of the purported benefits of MR is the ability to study lifetime effects or the effect of having a higher or lower exposure level over the entire lifetime. Though the age range in the UK Biobank limited us to explore at most a 25-year exposure window, the increase in bias with exposure window length suggests that biases may be even larger when studying lifetime effects with even longer exposure windows.

The biases when the outcome was simulated at age 65 was larger than that at the observed age. It is important to note that the exposure windows used in these scenarios are extremely different in nature. The exposure windows at age 65 imply that changes in the magnitude of the effect of the exposure are only due to age whereas the exposure windows at the age at baseline imply that changes in the effect of exposures are only due to time since exposure and are independent of age. Therefore, these results do not suggest that bias due to this phenomenon will be smaller when using cohorts with participants that have wide ranges in age. It suggests only that biases may be lower with exposure windows that are independent of age. Even in this scenario, however, biases tended to increase with the length of the exposure windows and relatively small biases such as the for the LDL cholesterol risk score may still be quite large when exposures truly have lifetime effects. The shape of the exposure also had an important impact on bias. In practice, many more exposure windows are possible.

One important aspect about the bias due to age-varying genetic associations is that it is dependent on the true exposure window during which the exposure has a causal effect on the outcome. This is important because the true exposure window is almost always unknown making it difficult to quantify the potential for this bias in practice. It is, however, possible to infer the direction of the bias in cases where the change in the genetic variant-exposure association changes only in one direction (*i*.*e*. either always increasing or always decreasing). When the genetic variant-exposure association continuously weakens with age, as was found with most genetics variants used as potential instruments for LDL cholesterol, the resulting bias must necessarily be positive (*i*.*e*. MR over-estimates the effect).

The UKBB are cross-sectional in nature meaning we could not actually follow people longitudinally to determine unequivocally how genetic associations change with age. Cohort effects may also contribute to the observed patterns. If cohort effects were entirely responsible for the differences in genetic associations across participants of different ages (i.e. the genetic associations are constant with age but differ across birth cohorts), this, on its own, would not bias MR estimates. If the causal effect of the exposure in question varied with age, however, the MR estimate would become a complier effect where the effect in birth cohorts with higher genetic associations would receive more weight. [Note: I removed mention of period affects here because, as far as I can imagine, they wouldn’t can the genetic associations to vary with age since, by definition, they should affect all ages equally]

The UK Biobank had a very low participation rate.^15^ It is possible that if participation was differential by genetic variant, exposure and age, that selection bias could explain the change in association by age. If selection bias were truly the explanation for this variation, then genetic associations observed at younger ages would likely be closest to the true genetic association. This could possibly entail even larger biases than that which we have observed here.

The exposures selected for this analysis are not intended to be representative of exposures used in MR. Therefore, we cannot say anything more generally about the potential for bias in MR analyses due to age-varying genetic associations. However, we have demonstrated the potential for important bias and therefore recommend that future MR studies use the tools developed for and supplied with this article to avoid using genetic variants whose effects change importantly over time or to inform themselves about the potential for bias.

These results indicate that, to understand both previously conducted MR studies and to better conduct MR studies in the future, investigators should carefully examine how the association between their proposed instruments and exposures vary over time. Future MR guidelines should also encourage MR studies to check this when possible, either in their own data, via literature review or external data for all genetic variant-exposure pairs. In data sets with participants that vary widely enough with age, this could be checked by modeling the interaction between age and number of copies of a genetic variant and plotting the results (a function to do this is included in the package associated with this paper: https://github.com/jalabrecque/MRcheck). If there is an important change in the genetic association over age, caution should be taken when interpreting MR estimates obtained via conventional MR approaches as they can suffer from bias due to this age-varying genetic association. When access to the data set is not possible (*e*.*g*. two sample MR) or there is not sufficient variation in age, quantifying bias from age-varying genetic associations will not be possible unless the proposed instruments have previously been studied in different age groups or genome-wide association study stratified by age. Though in some cases the direction of the bias is known, the current study and prior work^3^ make clear that statements about the magnitude of the bias require strong assumptions about the shape and length of the exposure window.

## Data Availability

The data are available from the UK Biobank to individuals who apply for and receive permission from the appropriate UK Biobank study committees.

