## Supplementary Material for "Age-varying genetic associations and implications for bias in Mendelian randomization"

Table 1: Table with the number of observations in each analysis, the average per allele effects ignoring age, and the average difference between difference numbers of copies of the genetics variant.

| | | | Per allele | | 1 vs 0 | | 2 vs 0 | |
|---|---|---|---|---|---|---|---|---|
| SNP | n | Effect/reference | est | se | est | se | est | se |
| **BMI** | | | | | | | | |
| rs1558902 | 354500 | A/T | -0.36 | 0.01 | -0.48 | 0.02 | -0.76 | 0.02 |
| rs6567160 | 354500 | C/T | -0.25 | 0.01 | -0.29 | 0.04 | -0.53 | 0.04 |
| rs13021737 | 354500 | G/A | -0.25 | 0.01 | -0.26 | 0.02 | -0.45 | 0.05 |
| rs10938397 | 354500 | G/A | -0.15 | 0.01 | -0.19 | 0.02 | -0.3 | 0.02 |
| rs543874 | 354500 | G/A | -0.23 | 0.01 | -0.25 | 0.04 | -0.48 | 0.04 |
| rs2207139 | 354500 | G/A | -0.2 | 0.02 | -0.22 | 0.05 | -0.42 | 0.05 |
| rs11030104 | 354500 | A/G | 0.19 | 0.01 | 0.16 | 0.04 | 0.36 | 0.04 |
| rs3101336 | 354500 | C/T | -0.11 | 0.01 | -0.1 | 0.02 | -0.22 | 0.02 |
| rs7138803 | 354500 | A/G | -0.13 | 0.01 | -0.17 | 0.02 | -0.27 | 0.03 |
| rs10182181 | 354500 | G/A | -0.17 | 0.01 | -0.21 | 0.02 | -0.34 | 0.02 |
| **Alcohol** | | | | | | | | |
| rs1229984 | 250736 | C/T | -2.01 | 0.1 | -2.06 | 0.1 | -2.62 | 0.75 |
| rs698 | 250736 | G/A | -0.2 | 0.03 | -0.22 | 0.06 | -0.4 | 0.06 |
| **C-reactive protein** | | | | | | | | |
| rs1130864 | 338337 | T/C | -0.27 | 0.01 | -0.29 | 0.03 | -0.55 | 0.03 |
| rs1205 | 338337 | C/T | 0.38 | 0.01 | 0.39 | 0.03 | 0.76 | 0.03 |
| rs3093077 | 338337 | G/T | -0.53 | 0.02 | -0.36 | 0.13 | -0.9 | 0.13 |
| **LDL cholesterol** | | | | | | | | |
| rs6511720 | 338436 | G/T | 0.16 | 0 | 0.15 | 0.01 | 0.31 | 0.01 |
| rs629301 | 338436 | T/G | -0.1 | 0 | -0.11 | 0 | -0.2 | 0.01 |
| rs1367117 | 338436 | A/G | -0.07 | 0 | -0.07 | 0 | -0.14 | 0 |
| rs4420638 | 338436 | G/A | -0.15 | 0 | -0.11 | 0.01 | -0.27 | 0.01 |
| rs4299376 | 338436 | G/T | 0.05 | 0 | 0.05 | 0 | 0.09 | 0.01 |
| rs2479409 | 338436 | G/A | 0.03 | 0 | 0.03 | 0 | 0.06 | 0 |
| rs1564348 | 338436 | C/T | -0.03 | 0 | -0.04 | 0.01 | -0.07 | 0.01 |
| rs11220462 | 338436 | A/G | -0.04 | 0 | -0.04 | 0.01 | -0.07 | 0.01 |
| rs6029526 | 338436 | A/T | -0.02 | 0 | -0.02 | 0 | -0.04 | 0 |
| rs635634 | 338436 | G/A | -0.03 | 0 | -0.04 | 0.01 | -0.07 | 0.01 |

**Table of associations between genetic variants and exposures**

**UK Biobank variable numbers**

BMI: 21001-0.0 Low density lipoprotein: f.307780 C-reactive protein: f.30710.0.0 Red wine: 1568-0.0
White wine: 1578-0.0
Beer: 1588-0.0
Spirits: 1598-0.0
Fortified wine: 1608-0.0 Age: 21022-0.0 Genetic principal components 1-10: 22009-0.1 - 22009-0.10

**Plots from Figure 2 for all genetic variants and polygenic risk scores.**

For each genetic variant, the left plot shows the relationship between age and the exposure in question for each genotype. The right plot is the per allele effect by age. For BMI and LDL the last plot is the association between the polygenic risk score and the exposure.

*BMI*

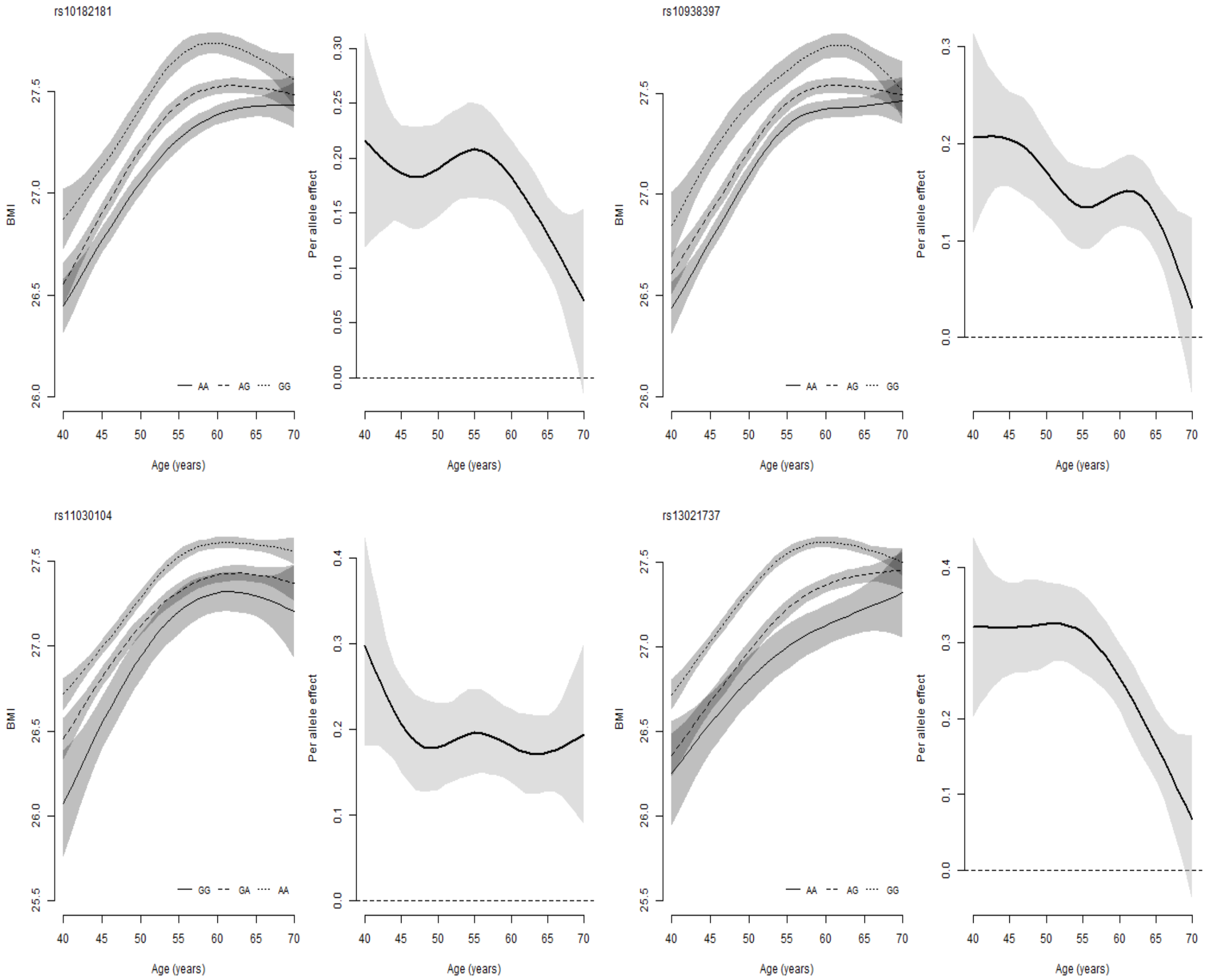

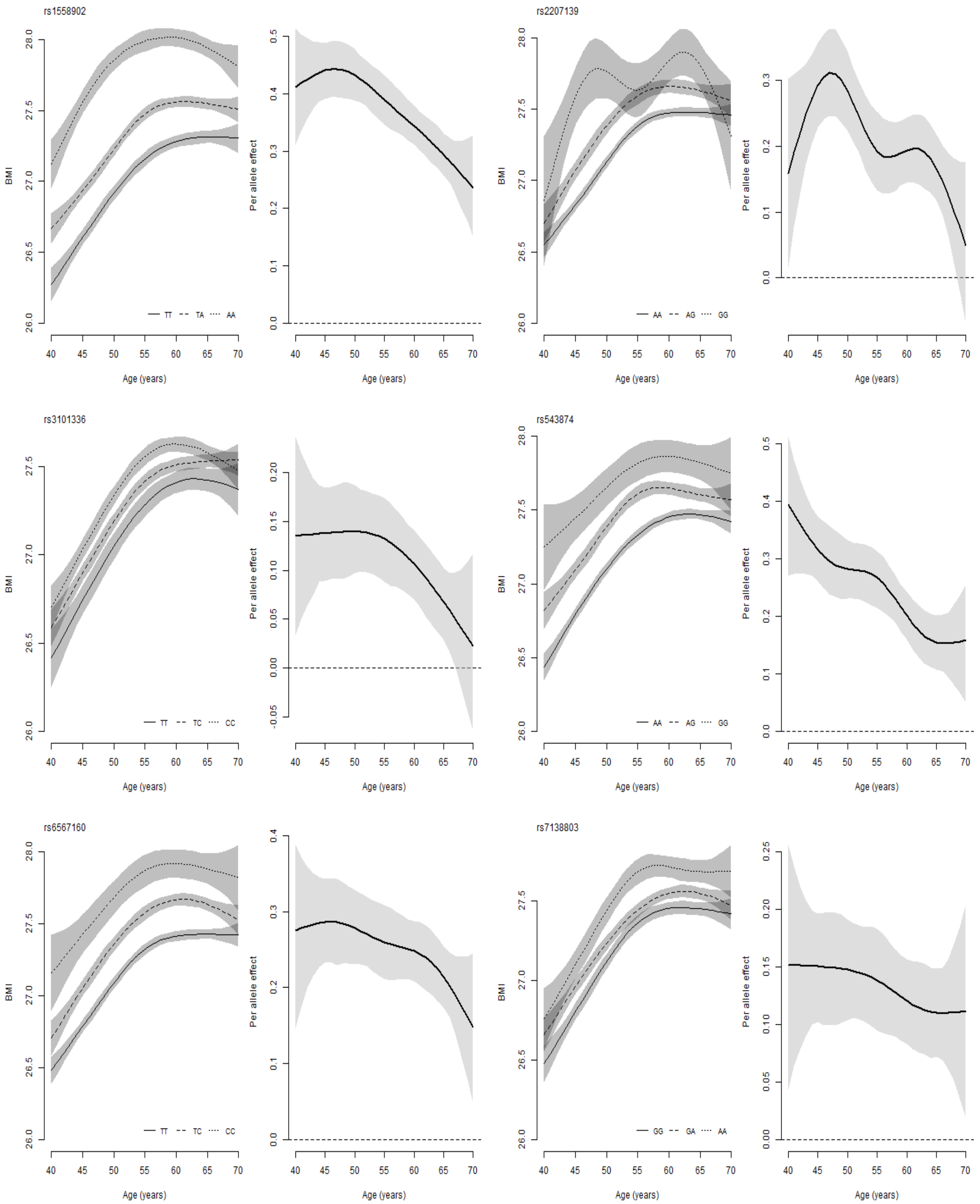

rs1558902
BMI
Age (years)
TT
TA
AA
Per allele effect
rs2207139
AA
AG
GG
rs3101336
TT
TC
CC
rs543874
AA
AG
GG
rs6567160
TT
TC
CC
rs7138803
GG
GA
AA

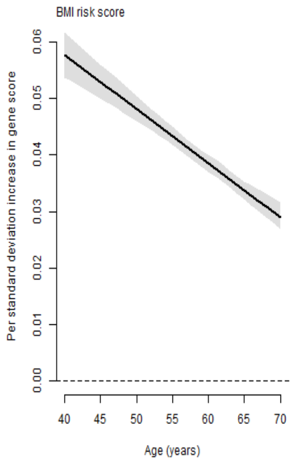
BMI risk score
Per standard deviation increase in gene score
0.00
0.01
0.02
0.03
0.04
0.05
0.06
40
45
50
55
60
65
70
Age (years)

*Alcohol consumption*

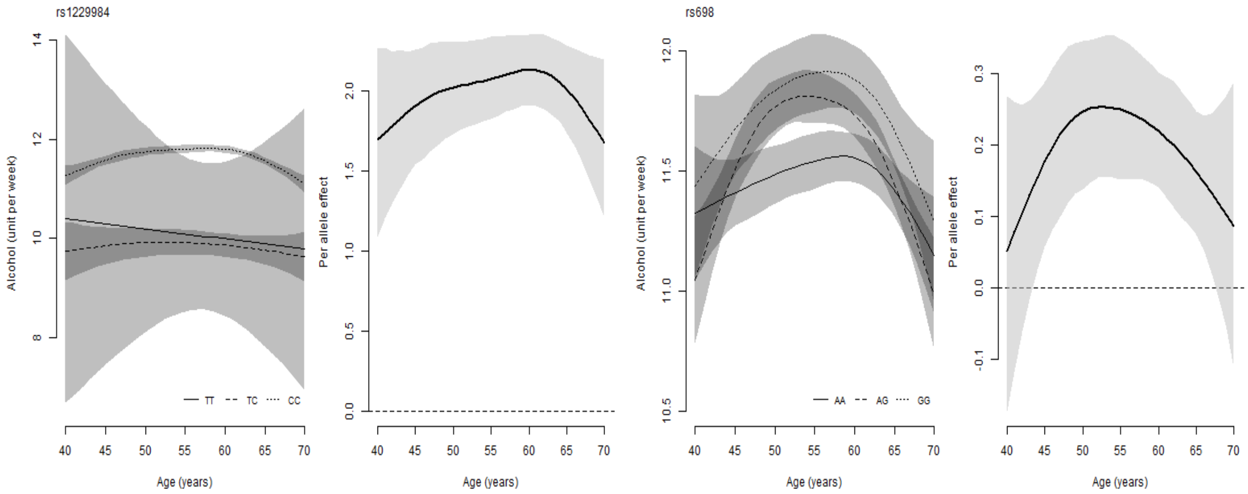

*C-reactive protein*

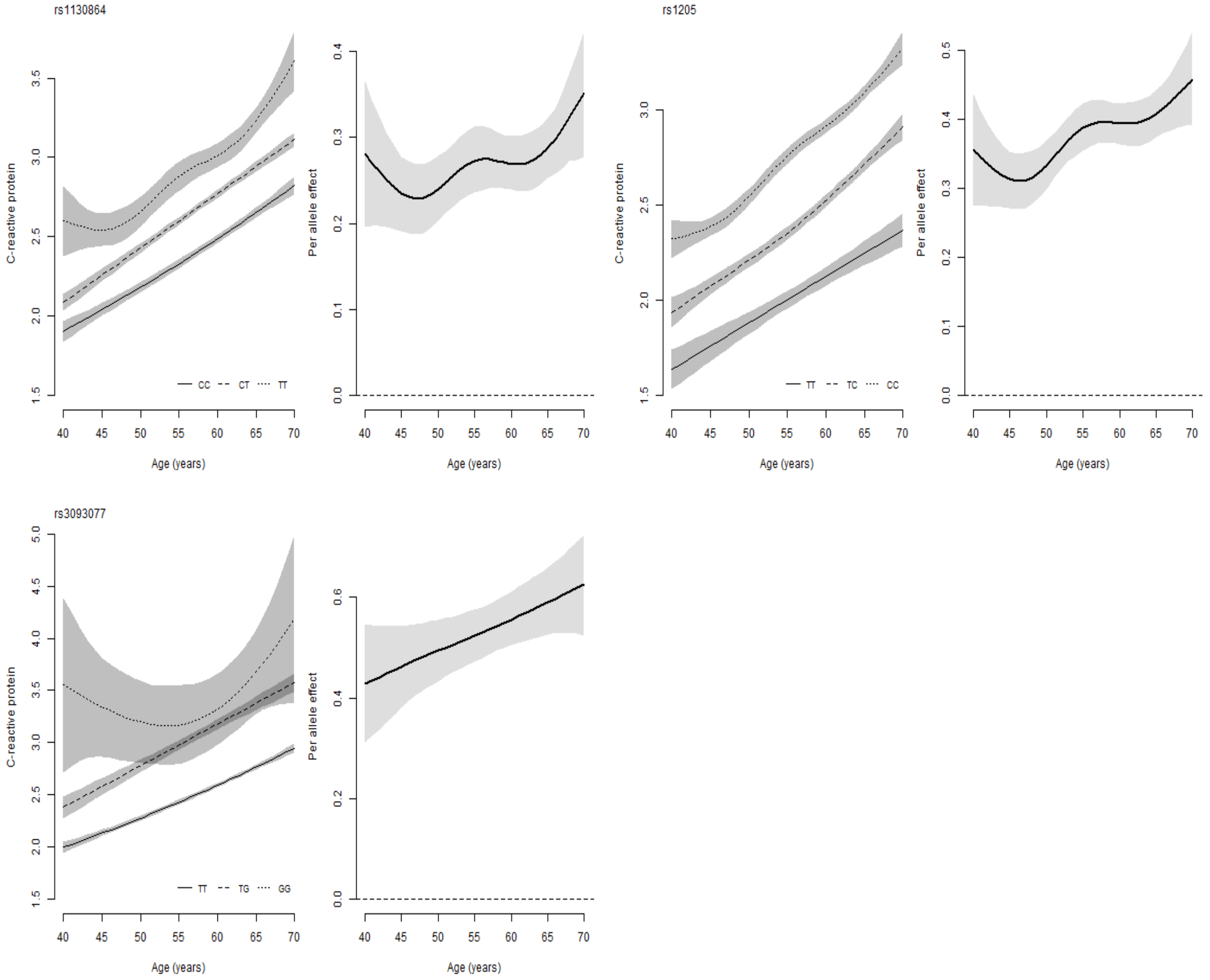

*LDL*

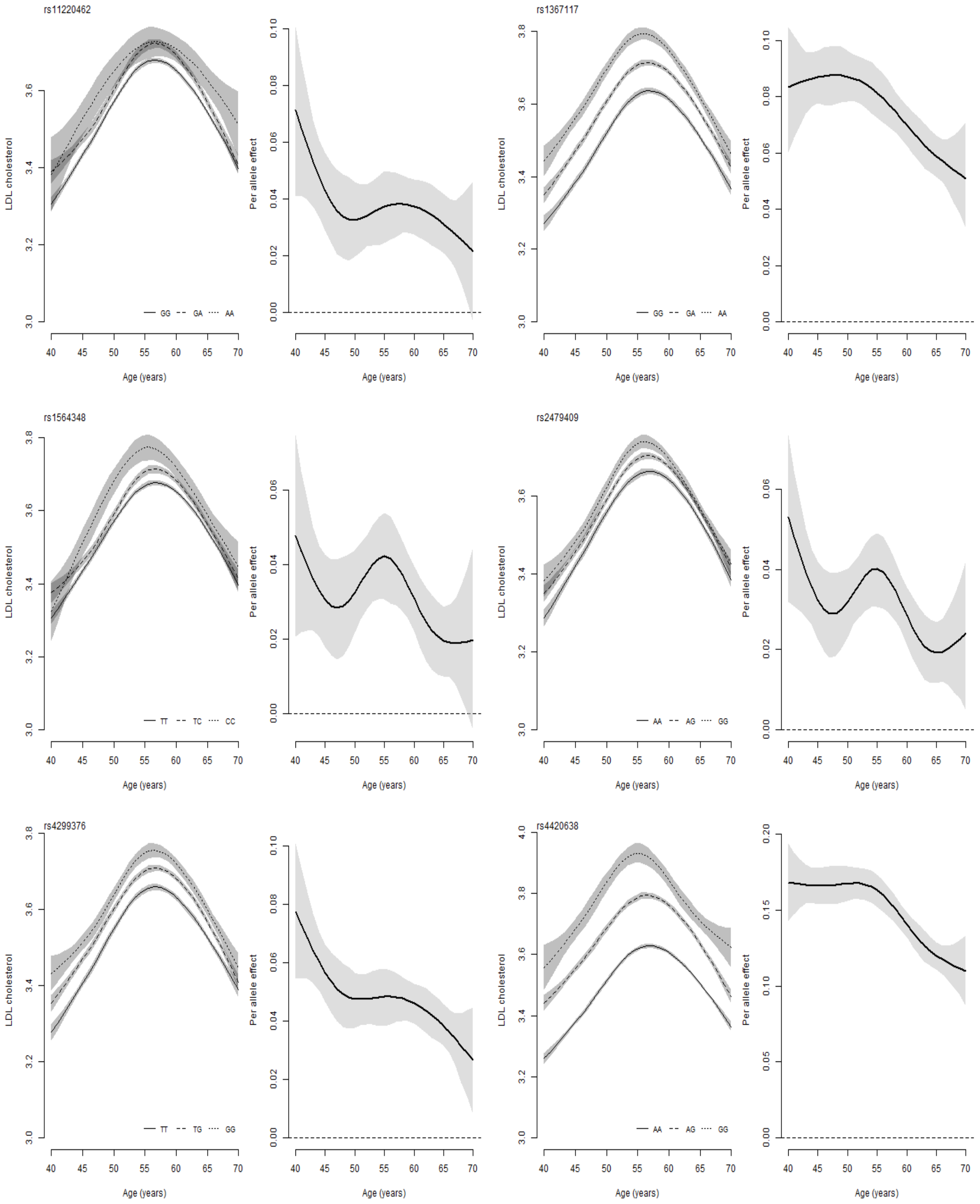

rs11220462
rs1367117
rs1564348
rs2479409
rs4299376
rs4420638
LDL cholesterol
Per allele effect
Age (years)
GG GA AA
TT TC CC
AA AG GG
TT TG GG

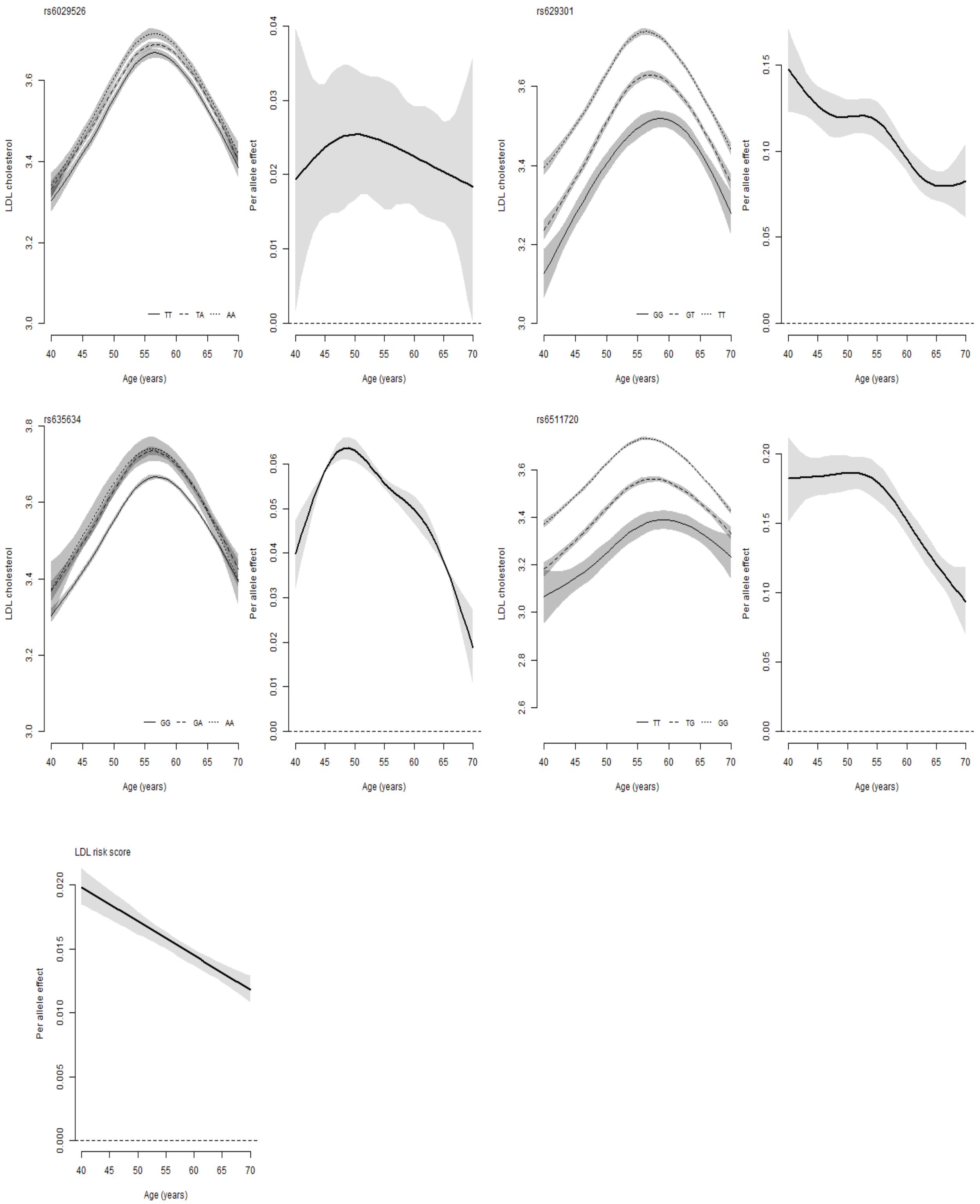
rs6029526
LDL cholesterol
Per allele effect
Age (years)
TT TA AA
rs629301
GG GT TT
rs635634
GG GA AA
rs6511720
TT TG GG
LDL risk score

### Code

*Data set-up*

```
library(magrittr)
library(plyr)
library(dplyr)

# Load dataset1
load("ds1.Rdata")

# Relatedness
rel <- read.table("UKB_relatedness.txt", header = TRUE)[,1:2]
names(rel) <- c("eid_gen_data","unrelated")
ds1 <- join(ds1, rel[,c("eid_gen_Data","unrelated")], by="eid__gen_data")
rm("rel")

# Exclusions and inclusions
ds1$include <- ds1$`21000-0.0`==1001 & ds1$unrelated==1

fileconn <- file("exclusions.txt")
writeLines(c("DESCRIPTIVES FOR EXCLUSIONS \n \n"),fileconn)
close(fileconn)

sink("exclusions.txt", append = TRUE)
cat("Table, white British:")
table(ds1$`21000-0.0`==1001)

cat("\n Table, unrelated:")
table(ds1$unrelated==1,useNA = "always")

cat("\n Table, unrelated and white British: \n ")
table(unrelated=ds1$unrelated==1,write_British=ds1$`21000-0.0`==1001,useNA = "always")

sink()

# Select and rename variables
ds1 %<>% filter(include==1) %>%
  select(eid,eid_gen_data,`21001-0.0`, `1498-0.0`, `21003-0.0`, `21022-0.0`,
                   `20116-0.0`, `20160-0.0`, `f.30710.0.0`, `f.30780.0.0`, `6138-0.0`,
                   contains("22009")) %>%
  rename(bmi = `21001-0.0`,
         age_assessment_centre = `21003-0.0`,
         age = `21022-0.0`,
         PC1 = `22009-0.1`,
         PC2 = `22009-0.2`,
         PC3 = `22009-0.3`,
         PC4 = `22009-0.4`,
         PC5 = `22009-0.5`,
         PC6 = `22009-0.6`,
         PC7 = `22009-0.7`,
         PC8 = `22009-0.8`,
```

```
         PC9 = `22009-0.9`,
         PC10 = `22009-0.10`,
         PC11 = `22009-0.11`,
         PC12 = `22009-0.12`,
         PC13 = `22009-0.13`,
         PC14 = `22009-0.14`,
         PC15 = `22009-0.15`,
         PC16 = `22009-0.16`,
         PC17 = `22009-0.17`,
         PC18 = `22009-0.18`,
         PC19 = `22009-0.19`,
         PC20 = `22009-0.20`,
         PC21 = `22009-0.21`,
         PC22 = `22009-0.22`,
         PC23 = `22009-0.23`,
         PC24 = `22009-0.24`,
         PC25 = `22009-0.25`,
         PC26 = `22009-0.26`,
         PC27 = `22009-0.27`,
         PC28 = `22009-0.28`,
         PC29 = `22009-0.29`,
         PC30 = `22009-0.30`,
         PC31 = `22009-0.31`,
         PC32 = `22009-0.32`,
         PC33 = `22009-0.33`,
         PC34 = `22009-0.34`,
         PC35 = `22009-0.35`,
         PC36 = `22009-0.36`,
         PC37 = `22009-0.37`,
         PC38 = `22009-0.38`,
         PC39 = `22009-0.39`,
         PC40 = `22009-0.40`,
         IDs = eid_gen_data) %>%
  filter(age>39 & age<71) # Filter age

# Load dataset2
load("ds2.Rdata")
ds2 %<>% rename(eid = `f.eid`,
                     crp = `f.30710.0.0`,
                     ldl = `f.30780.0.0`) %>%
  select(-contains("f.3"))

# Load dataset3
load("ds3.Rdata")

ds3 %<>% select(eid,`f.20117.0.0`, `f.1558.0.0`, `f.1568.0.0`,
                     `f.1578.0.0`, `f.1588.0.0`, `f.1598.0.0`,
                     `f.1608.0.0`) %>%
  rename(alc_status = `f.20117.0.0`,
         alc_freq = `f.1558.0.0`,
```

```r
         redwine = `f.1568.0.0`,
         whitewine = `f.1578.0.0`,
         beer = `f.1588.0.0`,
         spirits = `f.1598.0.0`,
         fortifiedwine = `f.1608.0.0`,
  ) %>%
  mutate(redwine = replace(redwine,redwine %in% c(-1,-3),NA),
         whitewine = replace(whitewine,whitewine %in% c(-1,-3),NA),
         beer = replace(beer,beer %in% c(-1,-3),NA),
         spirits = replace(spirits,spirits %in% c(-1,-3),NA),
         fortifiedwine = replace(fortifiedwine,fortifiedwine %in% c(-1,-3),NA),
         alc_units = whitewine + redwine + beer + spirits + fortifiedwine)

ds <- join(ds1, ds2, by = "eid") %>%
  join(ds3, by = "eid")
rm(list = c("ds1", "ds2", "ds3"))

# Load genetic data
library(data.table)
gen <- fread("genetic_data.txt.gz", header = TRUE)
gen <- data.frame(gen$IDs,
                  lapply(gen[,-1], function(x) {
  return(as.factor(round(x,0)))
})
) %>%
  rename(IDs = "gen.IDs")
ds %<>% plyr::join(., gen, by = "IDs")
rm("gen")

## Genetic data for last LDL snp
ldl_snp <- read.table("SNP_extra.txt", header=TRUE)
ldl_snp$rs635634.rs635634 <- as.factor(round(ldl_snp$rs635634.rs635634))
ds %<>% plyr::join(.,ldl_snp)

# Set up PRS for BMI
betas <- read.csv("betas_BMI.csv")

bmi_b <- betas[betas$phenotype=="BMI",]
bmi_b$beta %<>% as.numeric
bmi_r <- runs[runs$phenotype=="BMI",]

q <- ds[,bmi_r$rs_varname[match(bmi_b$SNP,bmi_r$rs)]]
rec_q <- apply(q,2,FUN= function(x) {
  2-as.numeric(x)
})
rec_q[,"rs11030104.11.27684517_A_G"] <-q$rs11030104.11.27684517_A_G

ds$bmi_score <- rec_q %*% bmi_b$beta
```

```r
# Set up PRS for LDL
ldl_betas <- read.csv("betas_ldl.csv")

ldl_betas$beta %<>% as.numeric
ldl_r <- runs[runs$phenotype=="LDL",]

q <- ds[,ldl_r$rs_varname[match(ldl_betas$rs,ldl_r$rs)]]
rec_q <- apply(q,2,FUN= function(x) {
  2-as.numeric(x)
})

ds$ldl_score <- rec_q %*% ldl_betas$beta

save(ds, runs, file="main_ds.Rdata")
```

*Data analysis: Individual genetic variants*

```r
library(devtools)
# install_github("https://github.com/jalabrecque/MRchecks")
library(MRchecks)

#
covars <- paste0("PC",1:10)
runs <- read.csv("SNP_exposure_combinations.csv")

# 1) Set up data ===============================================================

# Load data
source("create_dataset.R")

# 2) Analysis to get bias  =====================================================

library(parallel)
ptm <- proc.time()

set.seed(440302)
out <- mclapply(1:nrow(runs), function(i) {

  SNP <- runs$SNP[i]
  exposure <- runs$exposure[i]

  dat <- ds[complete.cases(select(ds,all_of(SNP),
                                  all_of(exposure),
                                  all_of(covars),
                                  age)),] %>%
    select(.,all_of(SNP),all_of(exposure),age,all_of(covars))

  mod <- do.call(SNPxAGE_model,
                 list(data = dat,
                      SNP = SNP,
                      phenotype = phenotype,
                      age = "age",
                      covars = covars,
                      k = 3))

  bias <- SNPxAGE_bias(mod, rep=1000)

  print(paste0("i=",i,"  ",proc.time()-ptm)[3])

  return(bias)

},mc.cores = 5) %>%
  set_names(paste0(runs$exposure,"_",runs$SNP))

print(proc.time()-ptm)
```

```r
save(out, file = "results_bias.Rdata")

# 3) Analysis to get per allele effect ==========================================

set.seed(440302)
out <- mclapply(1:nrow(runs), function(i) {

  SNP <- runs$SNP[i]
  exposure <- runs$exposure[i]

  dat <- ds[complete.cases(select(ds,all_of(SNP),
                                  all_of(exposure),
                                  all_of(covars),
                                  age)),] %>%
    select(.,all_of(SNP),all_of(exposure),age,all_of(covars))

  mod <- do.call(SNPxAGE_model,
                 list(data = dat,
                      SNP = SNP,
                      phenotype = phenotype,
                      age = "age",
                      covars = covars,
                      k = 3))

  return(SNPxAGE_effect(mod,reps = reps, plot=FALSE))

},mc.cores = 5) %>%
  set_names(paste0(runs$exposure,"_",runs$SNP))

print(proc.time()-ptm)

save(out, file = "results_per_allele.Rdata")
```

```
SNPxAGE_model_score <- function(data, score, phenotype, age, covars, k=3, indices=NULL,
                    pred_ages = NULL, type = "cr") {

  library(mgcv)
  library(magrittr)
  library(dplyr)

  # Run model -----------------------

  ## Assign age and SNP variables
  data$score <- data[,score]
  data$age <- data[,age]

  ## Run model

  mod <- mgcv::bam(formula = reformulate(termlabels = c("score*age",covars),
                                         response = phenotype),
                   data = data,
                   family = fam)

  mod$data <- data

  return(list(model = mod,
              params = as.list(match.call())))

}

SNPxAGE_bias_score <- function(SNPxAGE_output, rep = 1000) {

  # Set up------------------
  ## Set up variables
  model <- SNPxAGE_output$model
  data <- model$data
  covars <- eval(SNPxAGE_output$params$covars)

  # Predict BMI for all ages adding in variation to give it a similar variation
  # to the original data

  sampled_params <- rmvn(rep,coef(model),model$Vp) ## 1000 replicate param. vectors

  # Exposure windows at age 65 (number = width of window in years, l = linear, g = gaussian)
  y_window_5l <- seq(0,1, length.out = 5 + 1)[-1]
  y_window_10l <- seq(0,1, length.out = 10 + 1)[-1]
  y_window_25l <- seq(0,1, length.out = 25 + 1)[-1]
  y_window_5g <- dnorm(-20:5,mean = 2.5, sd = 1.2473)
  y_window_10g <- dnorm(-15:10,mean = 5, sd = 2.734)
  y_window_25g <- dnorm(0:25,mean = 12.5, sd = 6.3775)
```

```
# Apply over the 1000 sampled sets of parameters
out_sam <- lapply(1:rep, FUN = function(set_params) {

  d_rep <- cbind(data, setNames(
    # Apply over ages
    lapply(40:65, function(x) {

      Xp <- predict(model,transform(data, age = x),type="lpmatrix", newdata.guaranteed = TRUE)

      return(Xp %*% sampled_params[set_params,])
    }),
    paste0("pred_",40:65))
  )

  # Applying exposure windows at age 65
  d_rep$y5 <- (as.matrix(select(d_rep,paste0("pred_",61:65))) %*% y_window_5l)/3
  d_rep$y10 <- (as.matrix(select(d_rep,paste0("pred_",56:65))) %*% y_window_10l)/5.5
  d_rep$y25 <- (as.matrix(select(d_rep,paste0("pred_",41:65))) %*% y_window_25l)/13
  d_rep$y5_gauss <- (as.matrix(select(d_rep,paste0("pred_",40:65))) %*% y_window_5g)/0.993
  d_rep$y10_gauss <- (as.matrix(select(d_rep,paste0("pred_",40:65))) %*% y_window_10g)/0.978
  d_rep$y25_gauss <- (as.matrix(select(d_rep,paste0("pred_",40:65))) %*% y_window_25g)/0.9573

  # Applying exposure windows at observed age
  d_rep_obs <- lapply(unique(d_rep$age)[unique(d_rep$age)<66], function(a) {

    ds_temp <- d_rep[d_rep$age==a,]

    if (a < 44 & a < 49) {
      ds_temp$y5_obs <- NA
      ds_temp$y10_obs <- NA
      ds_temp$y5_gauss_obs <- NA
      ds_temp$y10_gauss_obs <- NA
    } else if (a < 49 & a >=44) {
      ds_temp$y5_obs <- (as.matrix(select(ds_temp,paste0("pred_",c((a-4):a)))) %*% y_window_5l)/3
      ds_temp$y10_obs <- NA
      ds_temp$y5_gauss_obs <- (as.matrix(select(ds_temp,paste0("pred_",c((a-4):a)))) %*% y_window_5g[
      ds_temp$y10_gauss_obs <- NA
    } else {
      ds_temp$y5_obs <- (as.matrix(select(ds_temp,paste0("pred_",c((a-4):a)))) %*% y_window_5l)/3
      ds_temp$y10_obs <- (as.matrix(select(ds_temp,paste0("pred_",c((a-9):a)))) %*% y_window_10l)/5.5
      ds_temp$y5_gauss_obs <- (as.matrix(select(ds_temp,paste0("pred_",c((a-4):a)))) %*% y_window_5g[
      ds_temp$y10_gauss_obs <- (as.matrix(select(ds_temp,paste0("pred_",c((a-9):a)))) %*% y_window_10
    }

    ds_temp$pred_obs <- ds_temp[,paste0("pred_",a)]

    return(ds_temp)

  }) %>% do.call(rbind,.) %>% as.data.frame
```

```
  # Estimation==================================================================

  iv_den <- summary(lm(d_rep$pred_65 ~ d_rep$score))$coef["d_rep$score",c("Estimate")]
  iv_avg <- summary(lm(d_rep[,SNPxAGE_output$params$phenotype] ~ d_rep$score))$coef["d_rep$score",c("

  out <- lapply(c("y5","y10","y25","y5_gauss","y10_gauss","y25_gauss"), FUN = function(y) {
    summary(ivreg(as.formula(paste0(y," ~ ",
                                    "pred_65",
                                    " | score")),
                  data = d_rep))$coef["pred_65",c("Estimate")]
  }) %>%
    do.call(c,.) %>%
    c(iv_den, iv_avg, .) %>%
    set_names(c("iv_den65","iv_avg","y5","y10","y25","y5_gauss","y10_gauss","y25_gauss"))

  # OBserved age
  #d_rep_obs$SNP_no_factor <- as.numeric(d_rep_obs$SNP)
  iv_den_obs <- summary(lm(d_rep_obs$pred_obs ~ d_rep_obs$score))$coef["d_rep_obs$score",c("Estimate")

  out_obs <- lapply(c("y5_obs","y10_obs","y5_gauss_obs","y10_gauss_obs"), FUN = function(y) {
    summary(ivreg(as.formula(paste0(y," ~ ",
                                    "pred_obs",
                                    " | score")),
                  data = d_rep_obs))$coef["pred_obs",c("Estimate")]
  }) %>%
    do.call(c,.) %>%
    c(iv_den_obs, .) %>%
    set_names(c("iv_den_obs","y5_obs","y10_obs","y5_gauss_obs","y10_gauss_obs"))

  return(c(out,out_obs))

}) %>%
  do.call(rbind,.)

output <- apply(out_sam, 2, FUN = function(param) {
  return(c(mean(param),sqrt(var(param)), quantile(x = param, probs = c(0.025,0.975))))
}) %>%
  t %>%
  set_colnames(c("est","se","q025","q975")) %>%
  set_rownames(c("iv_den65","iv_avg","y5","y10","y25","y5_gauss","y10_gauss",
                 "y25_gauss","iv_den_obs","y5_obs","y10_obs","y5_gauss_obs","y10_gauss_obs"))

return(output)
```

```
}

SNPxAGE_effect_score <- function(SNPxAGE_output, reps=1000, ages=40:70, plot=FALSE) {
  model <- SNPxAGE_output$model
  data <- model$data
  data$SNP_no_factor <- as.numeric(as.character(data$score))
  covars <- eval(SNPxAGE_output$params$covars)

  sampled_params <- rmvn(reps,coef(model),model$Vp)

  des_mat <- model.matrix(~ SNP_no_factor, data = data)

  out <- lapply(ages, function(age_) {

    data$age <- age_

    Xp <- predict(model,data,type="lpmatrix", newdata.guaranteed = TRUE)

    rep_out <- sapply(1:reps, function(r_num) {

      data$pred_pheno <- Xp %*% sampled_params[r_num,]

      lm.fit(des_mat,data$pred_pheno)$coef["SNP_no_factor"]

    })
    #print(age_)
    return(rep_out)
  }) %>% do.call(rbind,.) %>%
    apply(., 1, function(col) {
      c(mean(col),sqrt(var(col)),quantile(col, probs = c(0.025,0.975)))
    }) %>%
    t %>%
    as.data.frame %>%
    set_colnames(c("est","se","q025","q975")) %>%
    set_rownames(paste0("age_",ages))

  if (plot) {

    ds <- out[,c("est","q025","q975")]
    y_range <- range(c(0,ds))

    if (y_range[abs(y_range)==max(abs(y_range))]<0) {
      ds <- -ds

      y_range <- range(c(0,ds))
    }

    plot(ages, ds$est , type = "n", xlim = c(40,70), ylim = y_range,
         ylab = paste0("Per allele effect on ",SNPxAGE_output$params$phenotype), xlab = "Age")
```

```
    abline(h=0, lty=2)

    polygon(c(ages, rev(ages)),
            c(ds$q025,rev(ds$q975)), col=adjustcolor("grey",alpha.f=0.5),
            border=NA)
    lines(ages,ds$est, type = "l", lwd = 2 )

  }

  return(out)
}
```

*Data analysis: Polygenic risk scores*

```
# 1) Set up data ===============================================================

# Load data
source("create_dataset.R")

# Parameters
#runs <- runs[runs$rs %in% c("rs1558902","rs1229984","rs3093077","rs6511720"),]
runs <- data.frame(score=c("bmi_score", "ldl_score"),
           exposure = c("bmi", "ldl"))

# 3) Analysis of bias ==========================================================

library(parallel)
ptm <- proc.time()

set.seed(440302)
out <- mclapply(1:nrow(runs), function(i) {

  score <- runs$score[i]
  exposure <- runs$exposure[i]

  dat <- ds[complete.cases(select(ds,all_of(score),
                                   all_of(exposure),
                                   all_of(covars),
                                   age)),] %>%
    select(.,all_of(score),all_of(exposure),age,all_of(covars))

  dat[,score] <- dat[,score]/sd(dat[,score])

  mod <- do.call(SNPxAGE_score,
                list(data = dat,
                     score = score,
                     phenotype = exposure,
                     age = "age",
                     covars = covars,
                     k = knots))

  return(SNPxAGE_bias_score(mod,reps = 1000, plot=FALSE))
```

```r
},mc.cores = 5) %>%
  set_names(paste0(runs$exposure,"_",runs$score))

save(out_ldlscore_bias, file = paste0("score_bias.Rdata"))

# 3) Analysis of per allele effect =============================================

library(parallel)
ptm <- proc.time()

set.seed(440302)
out <- mclapply(1:nrow(runs), function(i) {

  score <- runs$score[i]
  exposure <- runs$exposure[i]

  dat <- ds[complete.cases(select(ds,all_of(score),
                                  all_of(exposure),
                                  all_of(covars),
                                  age)),] %>%
    select(.,all_of(score),all_of(exposure),age,all_of(covars))

  dat[,score] <- dat[,score]/sd(dat[,score])

  mod <- do.call(SNPxAGE_score,
                 list(data = dat,
                      score = score,
                      phenotype = exposure,
                      age = "age",
                      covars = covars,
                      k = knots))

  return(SNPxAGE_effect_score(mod,reps = 1000, plot=FALSE))

},mc.cores = 5) %>%
  set_names(paste0(runs$var,"_",runs$rs))

print(proc.time()-ptm)

save(list = obj_name, file = "effects_scores.Rdata")
```
